# Functional Personalized Treatment of Metastatic Porocarcinoma

**DOI:** 10.1101/2023.05.09.23289625

**Authors:** Sharon Pei Yi Chan, Chen Ee Low, Chun En Yau, Tzu Ping Lin, Weining Wang, Sam Xin Xiu, Po Yin Tang, Baiwen Luo, Nur Fazlin Bte Mohamed Noor, Kristen Alexa Lee, Jianbang Chiang, Tan Boon Toh, Edward Kai-Hua Chow, Valerie Shiwen Yang

**Author notes:** Correspondence to: Dr. Valerie Shiwen Yang, Translational Precision Oncology Laboratory, Institute of Molecular and Cell Biology, A*STAR, Singapore 61 Biopolis Drive, Proteos, Singapore 138673. Contributed equally (Sharon Pei Yi Chan, Chen Ee Low, Chun En Yau, Tzu Ping Lin).

## Abstract

Metastatic porocarcinomas (PC) have high mortality rates of 70%. PC is extremely rare, accounting for 0.01% of malignant cutaneous neoplasms, precluding the possibility of conducting clinical trials to evaluate treatment. Pathogenesis and clinical management of PC remain largely unknown Surgical resection of localized PC remains the mainstay of treatment. There are no effective agents for unresectable PC. Comprehensive genomic profiling did not yield any actionable genomic aberrations in our patient with metastatic PC. *Ex vivo* drug testing predicted pazopanib efficacy and treatment elicited remarkable clinicoradiological response. A functional precision medicine approach may be effective for the treatment of rare cancers.

## Introduction

Porocarcinoma (PC) is a rare subtype of cutaneous adnexal carcinoma, originating from the intraepithelial portion of eccrine sweat glands.^1^ PC can present on the legs, head and neck with no specific predilection of site.^2^ Although rare, it has the ability to metastasize to other parts of the body with mortality rates of 60 to 70%.^3^ Previous epidemiological studies estimate PC to only account for 0.005 to 0.01% of all malignant cutaneous neoplasms.^4,5^ Due to its rarity and lack of studies, both the pathogenesis and clinical management of PC remain poorly understood.

PC has been thought to develop from pre-existing poromas.^6^ Studies have also proposed that exposure to sunlight^7^ or immunosuppression^8^ were the main risk factors. As for the genetic mechanisms behind PC tumorigenesis, specific oncogenic drivers, including cell cycle and signaling pathways have been implicated. These pathways involve tumor protein 53 (TP53), HRas proto-oncogene, GTPase (HRAS), retinoblastoma 1 (Rb1), cyclin-dependent kinase inhibitor 2A (CDKN2A), epidermal growth factor receptor (EGFR), phosphatidylinositol-4,5-biphosphate 3-kinase, AKT serine/threonine kinase (PI3K-AKT) and mitogen-activated protein kinase (MAPK).^3^

Surgical resection of the primary tumor remains the mainstay of treatment for localized PC and is performed if possible. However, there is no clear management guideline for unresectable PC. Chemotherapy has been attempted, despite insufficient evidence to support the use of any specific regimen. Most unresectable or metastatic patients reported in the literature who underwent chemotherapy were treated with platinum-containing drugs such as carboplatin and cisplatin, achieving minimal success.^3^ Radiotherapy (RT) is sometimes used in combination with chemotherapy or after surgery.^9^ Neither chemotherapy nor RT has yielded good outcomes. Therefore, more effective treatment strategies are needed.

Here, we present a functional personalized treatment approach to guide the treatment of a patient with metastatic PC, achieving remarkable clinicoradiological response.

## Methods

### Case presentation

A lady in her 50s first presented to an external institution with three-month history of an enlarging left hip lump in August 2019. She underwent primary resection with close pathological margins of 2 mm. Staging computed tomography (CT) scan post-surgery showed a suspicious 1.1 cm left inguinal lymph node, which was subsequently confirmed to be metastatic disease on fine needle aspiration. She was then referred to the surgical department at our institution and underwent a wider excision and left groin lymph node dissection in October 2019, two months after the primary resection. Histology of the left groin tumor revealed no residual carcinoma in the tumor bed, but 21 out of 25 lymph nodes showed macro-deposits of metastatic PC. Additional stains for estrogen receptor (ER), progesterone receptor (PR) and HER2 were negative. The patient was given adjuvant radiotherapy 50 Gray (Gy) over 25 sessions to the pelvis and left groin from December 2019 to February 2020. In February 2021, surveillance positron emission tomography and computed tomography (PET-CT) scans showed prominent left para-aortic, right common iliac, and left external lymph nodes with mild to moderate FDG-uptake, suggesting nodal disease recurrence. She was discussed at a multidisciplinary tumor board and decision was made for close interval CT scan in view of no incremental nodal disease, though the lesions were FDG-avid on PET-CT. CT scans of her thorax, abdomen, and pelvis performed three months later in May 2021 showed multiple new pulmonary nodules in the lungs, as well as enlarging retroperitoneal, pelvic, left supraclavicular and left axillary lymph nodes. She remained clinically well other than episodic dry cough and was placed on watchful waiting. Patient underwent a left supraclavicular lymph node biopsy which confirmed the recurrence of metastatic PC in November 2021. In January 2022, she developed worsening dyspnea and left lower limb swelling. Ultrasound of her deep veins did not indicate thrombosis. On examination, she was wheelchair-bound, on oxygen supplementation, and had left lower limb swelling up to her thigh.

### Pre-treatment evaluation

Formalin-fixed paraffin-embedded tumor tissue was sent for immunohistochemistry (IHC) for diagnostic evaluation. Oncomine Comprehensive Assay Plus was also performed. Dissociated cells from biopsies of her lung metastases were used for *ex vivo* drug testing.

### Ex vivo drug testing

Fresh biopsies of the patient’s lung metastases were digested with a GentleMACS dissociator in warm RPMI-1640 medium (Biowest) with 100 μg/mL LiberaseTM (Sigma Aldrich, Singapore) for five minutes and then incubated on a shaker at 70 rpm in a cell culture incubator for two hours. Following filtration and centrifugation, the isolated cells were collected, resuspended in an appropriate volume of RPMI-1640 medium supplemented with 20% (v/v) fetal bovine serum (FBS; Gibco, USA), 1% (v/v) penicillin/streptomycin (Gibco, USA) and seeded in 384-well plates at a density of 2000 cells/ well for 24 hours prior to treatment. After 48 hours of drug treatment at concentrations ranging from 0.000001 μM to 100 μM, cell viability was measured using CellTiter-Glo (CTG) Luminescent Cell Viability Assay (Promega) according to the manufacturer’s instructions. To compute IC_50_ values, sigmoidal dose-response curves were generated using Prism 9 software (GraphPad) by fitting quantified cell viability. The list of drugs used are shown in Figure 2A-C.

## Results

### Histology and Comprehensive Genomic Profiling

Sections of supraclavicular and groin lymph nodes revealed nests and islands of malignant epithelioid cells with ample eosinophilic cytoplasm and oval vesicular nuclei (Figure 1A-C). Additionally, there was suggestion of duct formation (Figure 1C). Overall histological features and prior history of PC confirm metastatic PC. Comprehensive genomic profiling (CGP) via Oncomine Comprehensive Assay Plus on her formalin-fixed paraffin-embedded tumour specimen demonstrated the existence of genomic alterations predominantly in three genes: KMT2D, MRE11A, TBX3 (Supplementary Materials). However, these alterations were not targetable.

**Figure 1:**
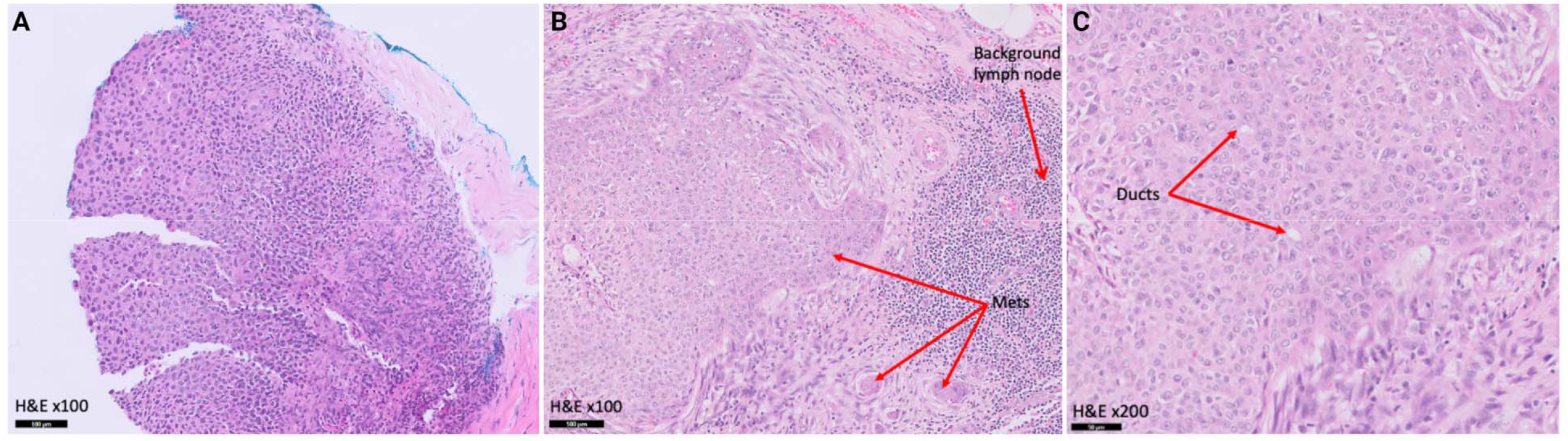
Hematoxylin & eosin (H&E) staining confirms the diagnosis of metastatic porocarcinoma to the lymph nodes. **Figure 1A:** H&E staining of supraclavicular lymph nodes showing porocarcinoma infiltration. Scale bar represents 100 μm. **Figure 1B:** H&E staining of groin lymph nodes showing metastasis. Scale bar represents 100 μm. **Figure 1C:** H&E staining of groin lymph nodes showing ducts formation. Scale bar represents 50 μm.

**Figure 2:**
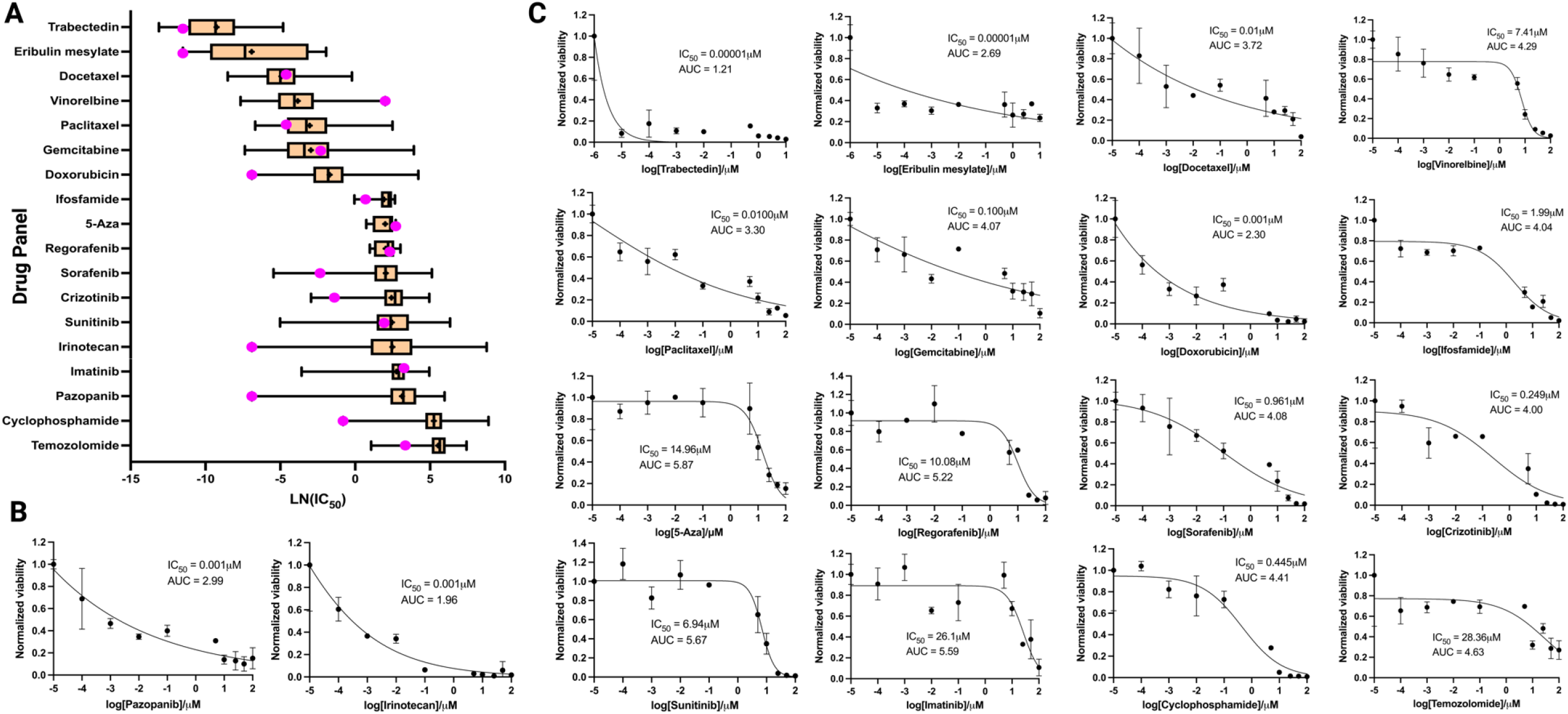
*Ex vivo* drug testing results of 18 different drugs on fresh biopsies of the patient’s lung metastases demonstrating efficacy of pazopanib. **Figure 2A:** Box plot comparing IC_50_ data with Genomics of Drug Sensitivity in Cancer (GDSC)^16^ dataset. Drugs used in our single-drug testing assay are listed on the y-axis and corresponding log transformed IC_50_ values listed on the x-axis. Boxes represent the 25^th^ to 75^th^ percentiles, and horizontal lines within the boxes represent the median values. ‘+’ represents mean IC_50_ values. The ends of the solid lines extending either side of the boxes represent the approximate 95% confidence intervals. **Figure 2B:** Dose-response curves for pazopanib and irinotecan the top two drugs with the largest leftward deviation of IC_50_ of the patient’s tumor sample from the mean IC_50_ values from the GDSC dataset. **Figure 2C:** Dose response curves of the remaining drugs in order of ascending IC_50_ values. All dose-response curves are represented as means (SD) of two technical replicates.

### Ex vivo drug testing

Compounds used consisted of a selected panel of FDA-approved agents that covers key cancer-associated signalling pathway targets including tyrosine kinases, TNF-alpha, DNMT, c-kit, mTOR, and ALK. Single-drug dose response assay was performed (Figure 1C) and individual IC_50_ values obtained were compared with the publicly available GDSC dataset (Figure 1A). The overall most cytotoxic drugs based on normalized dose responses were pazopanib (multi-tyrosine kinase inhibitor; IC_50_ 0.001μM) and irinotecan (Topoisomerase I inhibitor; IC_50_ 0.001μM) (Figure 1B). Both drugs displayed significantly lower IC_50_ values in the patient’s cells as compared to almost 1000 other human cancer cell lines within the GDSC resource, suggesting strong sensitivity of the patient’s tumor cells towards these compounds.

### Clinical response

Based on the lack of standard systemic treatment options in PC, the lack of actionable targets on CGP of the tumor sample, as well as *ex vivo* drug testing results, the patient was started on the oral multi-target tyrosine kinase inhibitor pazopanib 800 mg in January 2022. She was offered pazopanib instead of doxorubicin or irinotecan as she was deemed unfit for chemotherapy in view of her poor performance status. When the patient attended the next clinic session on February 2022, she demonstrated remarkable clinical improvement in both her dyspnea and lower limb swelling. She was able to walk upslope for up to 500 meters, without oxygen supplementation, support, or rest.

### Clinical investigations

In January 2022 prior to molecular profiling and *ex vivo* drug testing, computed tomography imaging of the thorax, abdomen and pelvis (CT TAP) revealed an increase in the number and size of numerous bilateral pulmonary nodules, suspicious for metastasis. The lymph nodes at the left axillary, supraclavicular, retroperitoneal and pelvic regions were enlarged and suspicious for nodal metastases. After two months of treatment with pazopanib, her CT TAP revealed a decrease in size and cavitation of multiple scattered bilateral pulmonary metastases. Right upper lobe lesion decreased from 23 mm to 20 mm (Figure 3A-B). Left upper lobe cavitary mass decreased from 42 mm to 34 mm (Figure 3A-B). The size of the other lymph nodes remained stable.

**Figure 3:**
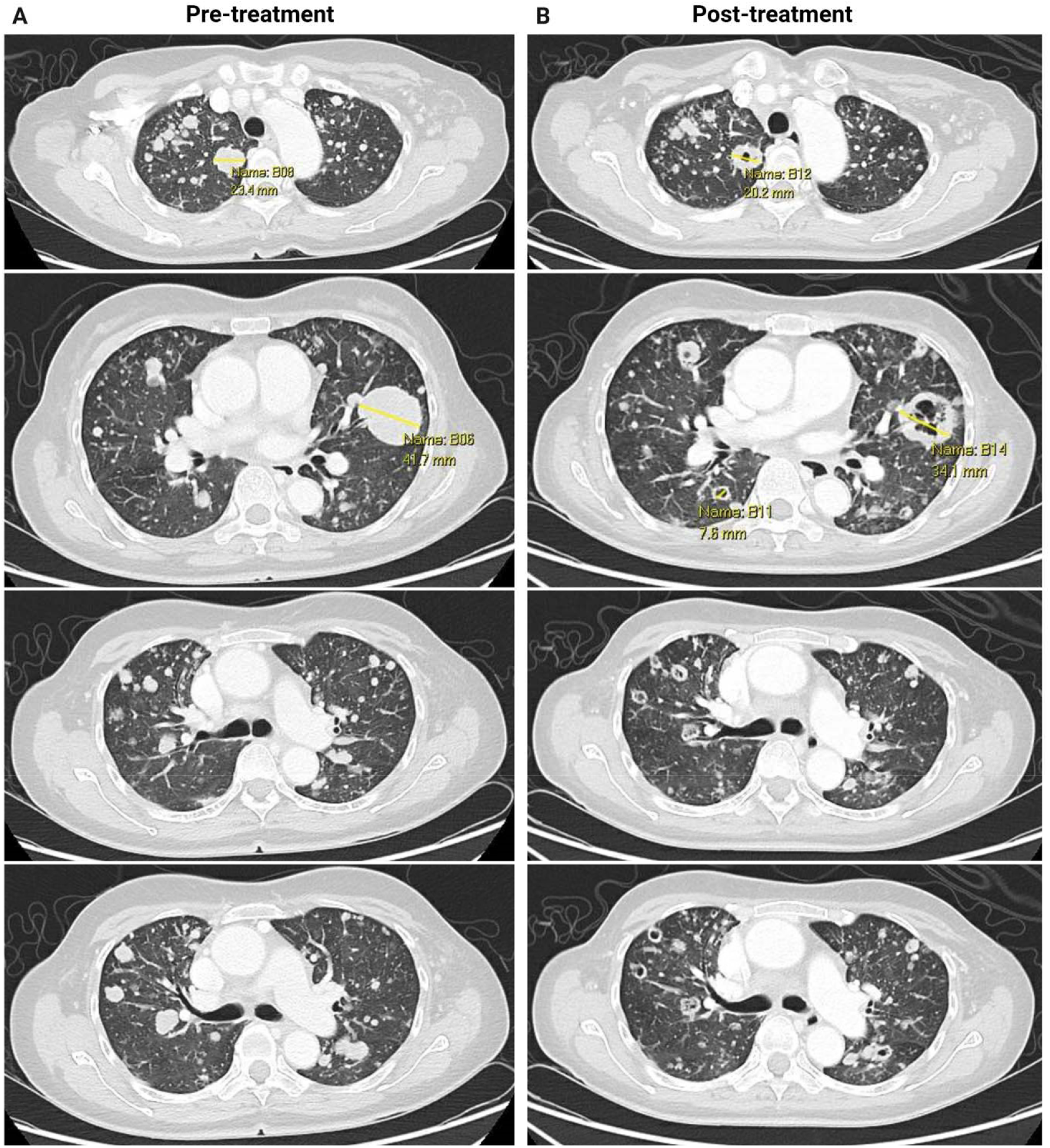
Computed tomography (CT) images of the chest before and after pazopanib treatment showing remarkable radiological outcomes. **Figure 3A:** CT images of the chest before pazopanib treatment in January 2022. **Figure 3B:** CT images of the chest after two months of pazopanib treatment in March 2022. Several bilateral pulmonary lesions demonstrating a decrease in size and cavitation. Cavitation suggests tumor liquefaction and necrosis, consistent with known patterns of radiological response to pazopanib. ^17^

### Outcome after treatment with pazopanib

The patient remained well on pazopanib for five months and did not report any drug-related side effects. When she was seen in the clinic in May 2022, she had right hip and back pain, without much relief with morphine. She was admitted for pain control and underwent CT TAP in June 2022. CT TAP showed stable disease in lymph nodes and lungs, but progression in her spine, with several compression fractures. As her performance status had improved with pazopanib treatment, she was offered a switch to palliative doxorubicin or irinotecan chemotherapy based on the *ex vivo* drug testing results, but she declined and opted for best supportive care. The patient eventually passed away on July 2022.

## Discussion

To the best of our knowledge, this is the first instance where a functional personalized treatment approach was used to guide the treatment of a patient with metastatic PC. We describe a patient with metastatic PC, for whom there is no known effective systemic treatment. To personalize a treatment approach for her deteriorating condition, we performed genomic profiling of her tumor tissue, of which no actionable findings were elucidated. We proceeded with *ex vivo* drug testing, revealing that pazopanib was one of the most potent yet tolerable drugs. The patient was started on pazopanib and achieved remarkable clinical improvement within a few weeks, maintaining disease control over five months before eventual disease progression.

Due to the rarity of this disease, there are no established management guidelines. Treatment information has been limited to case reports or series. Current treatment options for metastatic PC patients include surgery, RT, and systemic options including chemotherapy, immunotherapy, and targeted therapy with palliative intent. The use of immunotherapy was highlighted in a case report of a 67-year-old lady with metastatic PC by Lee *et al*.^10^ She was started on pembrolizumab against programmed death-ligand 1 after 12 cycles of carboplatin and capecitabine failed to stop disease progression. The patient achieved an excellent response. In another case report, targeted therapy against EGFR using cetuximab and paclitaxel has been shown by Godillot *et al*.^11^ to demonstrate clinical improvement in one patient. PET-CT showed a complete metabolic response and almost complete morphological response. However, both Lee *et al*. and Godillot *et al.* postulated that the positive therapeutic response is likely due to strong PD-L1 or EGFR expression respectively and may not be effective in patients without expression, as in the case of our patient.

This is the first patient where pazopanib has demonstrated its potency in treating metastatic PC. Pazopanib is an oral multi-tyrosine kinase inhibitor (TKI) that serves to inhibit intracellular TKI of vascular endothelial growth factor receptors −1, −2 and −3, platelet-derived growth factor receptors −α, and −β, leukocyte-specific protein tyrosine kinase, interleukin-2 receptor-inducible T-cell kinase, colony-stimulating factor-1 receptor, fibroblast growth factor receptors, and the stem-cell factor receptor c-KIT. Although pazopanib was developed for use against various cancers, it is currently only approved for the treatment of renal cell carcinoma and advanced soft-tissue sarcoma.^12^ While the tumorigenic pathways in PC affected by pazopanib are poorly understood, our patient managed to improve dramatically with pazopanib. Further studies should focus on uncovering the specific pathways responsible for pazopanib efficacy in PC or consider incorporating pazopanib into PC treatment regimens.

Fundamentally, this work gives credence to the use of functional personalized treatment. Precision oncology has traditionally utilized static features of tumors such as expression of key targets or genomic analysis to guide treatment.^13^ However, actionable targets are not often found and the efficacy of this approach has been underwhelming, with several precision oncology trials yielding low clinical benefit.^14,15^ Perhaps unsurprisingly, elucidating the static features of the tumor yielded three genomic targets, but they were not actionable. On the other hand, a functional personalized treatment approach identified pazopanib efficacy and eliciting a striking disease response in this patient. While further studies are required to unravel the pathways underlying this response, this supports the potential of using functional personalized treatment as an effective approach to treat rare cancers with no established treatment options. Future research should streamline how functional assays could be correlated to clinical responses, as well as limiting pre-analytical variability of viable samples.

## Conclusion

In summary, this patient is the first patient to undergo a functional personalized treatment for metastatic porocarcinoma, and the results offer proof-of-concept for such an approach in rare cancers with no systemic treatment options. Given the promising results, this approach should be tested and validated in a larger number of patients.

## Consent for publication

Consent for publication has been obtained from the patient.

## Data Availability

All data produced in the present study are available upon reasonable request to the authors.

