## Supplementary Materials for "Functional Personalized Treatment of Metastatic Porocarcinoma"

**Supplementary Table 1:** Oncomine Comprehensive Assay Plus results for possible targetable mutations and their frequency. These results indicated the presence of three non-actionable genomic alterations.

| **SNV & INDEL** | | | |
| --- | --- | --- | --- |
| **Gene** | **Variant Type** | **Genomic Alterations Identified** | **Variant Frequency** |
| KMT2D  (NM_003482) | SNV | Exon39, c.11791C>T  p.Leu3931Phe (p.L3931F) | 15.70% |
| MRE11A  (NM_005590) | SNV | Exon8, c.668A>G  p.His223Arg (p.H223R) | 43.90% |
| TBX3  (NM_005996) | DEL | Exon6, c.1432_1433delCT  p.Leu478fs (p.L478fs) | 23.20% |

**Supplementary Table 2:** Interpretation of Oncomine Comprehensive Assay results

| **GENE** | **INTERPRETATION** |
| --- | --- |
| KMT2D  missense_variant  Exon39 c.11791C>T p.Leu3931Phe (p.L3931F) | This variant indicates a C to T substitution at nucleotide 11791, leading to an amino acid substitution from Leucine to Phenylalanine at codon 3931. This variant has not been reported in earlier cases of cancers [1]. The functional effects of this variant have not been biochemically characterized. Thus, the clinical and functional significance of this variant remains unknown at this point in time.  KMT2D is a histone methyltransferase that methylates 'Lys-4' of histone H3 (H3K4me). H3K4me represents a specific tag for epigenetic transcriptional activation. KMT2D is also identified as a coactivator for estrogen receptor by being recruited by ESR1, thereby activating transcription. |
| MRE11A  missense_variant  Exon8  c.668A>G p.His223Arg (p.H223R) | This variant indicates a A to G substitution at nucleotide 668, leading to an amino acid substitution from Histidine to Arginine at codon 223. This variant has not been reported in earlier cases of cancers [1]. This effect of this variant has not been functionally characterized. Computational predictions suggest a pathogenic role of this variant (SIFT: damaging, PROVEAN: deleterious). The clinical and functional significance of this variant remains unknown at this point in time.  This gene is also known as MRE11. This gene encodes a nuclear protein involved in homologous recombination, telomere length maintenance, and DNA double-strand break repair. By itself, the protein has 3' to 5' exonuclease activity and endonuclease activity. The protein forms a complex with the RAD50 homolog; this complex is required for nonhomologous joining of DNA ends and possesses increased single-stranded DNA endonuclease and 3' to 5' exonuclease activities. Loss of MRE11 or inhibition of MRE11 can lead to hypersensitivity to genotoxic agents. The endonuclease and exonuclease function of MRE11 complicates its role in cancer. MRE11 expression confers different effects under distinct contexts, where its alteration may lead to effects ranging from predisposition to malignancy to inducing chemoresistance. |
| TBX3  frameshift_variant  Exon6  c.1432_1433delCT p.Leu478fs (p.L478fs) | This variant indicates the deletion of 2 bases between nucleotides 1432 and 1433, leading to a frameshift from Leucine at codon 478 onwards. This variant has not been reported in earlier cases of cancer. In general, frameshift mutations lead to a loss of function as it leads to the production of a truncated protein. This variant occurs within the activation domain of TBX3, potentially leading to the loss of the activation function of TBX3. The clinical and functional significance of this variant remains unknown.  TBX3 is a transcriptional repressor involved in developmental processes. It is hypothesized to play a role in limb pattern formation and acts as a negative regulator of PML function in cellular senescence. |
